# Cost-effectiveness of comparative survey designs for helminth control programs: post-hoc cost analysis and modelling of the Kenyan national school-based deworming program

**DOI:** 10.1101/2023.08.11.23293966

**Authors:** Mark Minnery, Collins Okoyo, Grace Morgan, Andrew Wang, Olatunji Johnson, Claudio Fronterre, Antonio Montresor, Suzy J. Campbell, Charles Mwandawiro, Peter Diggle

## Abstract

**Background:** Soil-transmitted helminths (STH) and schistosomiasis comprise the most wide-spread NTDs globally. Preventative chemotherapy is a cost-effective approach to controlling morbidity of both diseases, but relies on large scale surveys to determine and revise treatment frequency. Availability of detailed information on survey costs is limited despite recent methodological surveying innovations. We micro-costed a survey of STH and schistosomiasis in Kenya, and linked results to precision estimates of competing survey methods to compare cost-efficiency.

**Methods:** Costs from a 2017 Kenyan parasitological survey were retrospectively analyzed and extrapolated to explore marginal changes when altering survey size, defined by the number of schools sampled and the number of samples taken per school. Subsequent costs were applied to simulated precision estimates of model-based geostatistical (MBG) and traditional survey designs. Cost-precision was calculated for a range of survey sizes per method. Four traditional survey design scenarios, based around WHO guidelines, were selected to act as reference cases for calculating incremental cost-effectiveness ratios (ICERs) for MBG design.

**Findings:** MBG designed surveys showed improved cost-precision, particularly if optimizing number of schools against samples per school. MBG was found to be more cost-effective under 87 of 92 comparisons to reference cases. This comprised 14 situations where MBG was both cheaper and more precise, 42 which had cost saving with precision trade off (ICERs; $8,915-$344,932 per percentage precision lost); and 31 more precise with increased cost (ICERs; $426-$147,748 per percentage precision gained). The remaining 5 comparisons represented extremes of MBG simulated site selection, unlikely to be applied in practice.

**Interpretation:** Efficiency gains are possible for deworming surveys when considering cost alone, such as through minimizing sample or analysis costs. However further efficiency maximization is possible when designing surveys using MBG given its improved precision and ability to optimize the balance between number of schools and sample size per school.

**Research in context:** *Evidence before this study:* Soil-transmitted helminths (STH) and schistosomiasis are widespread neglected tropical diseases (NTDs) which require preventative chemotherapy (PC) for morbidity control among school–aged children. A key component of PC for both diseases is the use of large-scale surveys to determine prevalence in order to guide treatment frequency, or in the case of very low prevalence, measure potential resurgence. Given the need for a SAC population estimate, surveys represent a substantial proportion of helminth control program budgets. As prevalence of STH and schistosomiasis reduce globally, there is a need to understand costing components of these surveys to make best use of available resources. Recent innovations based on an increasingly sophisticated existing literature, in survey design using geospatial statistical methods to select survey sites have been shown to deliver more precise results, given the same resources, than traditional design approaches. To date, few studies have reported costs of large scale STH and schistosomiasis studies in sufficient detail to allow cost-effectiveness comparisons of geospatially designed surveys against traditional design approaches using real world data.

*Added value of this study:* Detailed bottom up costing is provided for a representative survey of STH and schistosomiasis conducted in Kenya in 2017. Results are analyzed and extrapolated to demonstrate how costs differ depending on numbers of schools surveyed and individuals sampled per school. Areas of potential efficiency maximization are highlighted. Costs are coupled with a previous simulation study comparing the precision of traditional and MBG-based design and analysis of the same series of surveys conducted in Kenya. A range of cost-precision estimates are generated and compared to show incremental cost-effectiveness ratios of both traditional and geospatial survey design under varying budget constraints, represented by survey size. The geospatial design is shown, under almost all reference case comparisons to be cost saving, more precise, or both.

*Implications of all the available evidence:* This study quantifies the potential increased efficiency that can be gained when geospatial methods are used to design and analyse large representative surveys of helminths. This is critical for the future of school-based deworming programs as a greater emphasis is placed on maintaining cost-effectiveness in environments where prevalence and morbidity due to STH and schistosomiasis are reducing.

## Introduction

Infections caused by soil-transmitted helminths (STH) and schistosomes comprise the two most wide-spread neglected tropical diseases (NTDs) globally with approximately 1.5 billion and 240 million people infected in 2021, respectively [1], [2]. Infection with both STHs and schistosomes can lead to local and systemic pathological effects including anaemia, growth stunting, impaired cognition, decreased physical fitness, and organ-specific effects [3][4], while severe cases can lead to intestinal obstructions and gangrene [4][5]–[7]. Both STH and schistosomiasis have been recognized as having significant developmental and educational effects on children which can hinder their ability to lead a full healthy life while also affecting productivity into adulthood [5].

Repeated preventative chemotherapy (PC) with albendazole or mebendazole for STH, and with praziquantel for schistosomiasis, is used to control helminth morbidity within at-risk populations [4]. All three drugs are well suited to PC given their known safety profile, tolerability and low cost, and are often administered through school-based deworming campaigns [8]. Using schools as a platform for PC allows a captive population for treatment, maintaining high coverage levels while minimizing cost and targeting those at most risk. Efficiency through this platform in combination with the long term benefits of PC, recently demonstrated by Hamory and colleagues [5], make school-based deworming a highly cost-effective intervention for child development [8], [9].

Treatment for STH and schistosomiasis is sufficiently inexpensive and safe to be warranted without prior diagnosis among individuals, provided there is an indication that infection is prevalent within the population [10], [11]. Population level prevalence is determined through large-scale surveys at repeated intervals during PC campaigns, currently recommended after every 3-5 rounds of PC that have achieved at least 75% treatment coverage among school age children (SAC) [10]. Surveys are used to adapt treatment to the current epidemiological situation. Guided by prevalence levels and recommendations from the WHO, PC frequency ranges from three times a year, to once every two years, with suspension recommended when prevalence is less than 2% [10]. Large-scale surveying is necessary to guide treatment frequency as helminth infections are not able to be systematically diagnosed through notifiable disease registers [12]. Given the need for population level estimates of SAC to determine PC frequency, the cost to determine prevalence and intensity of STH and schistosomiasis comprises a substantial proportion of overall school-based deworming budgets [7].

Greater consideration is needed to understand the costs involved in surveying and possible strategies for reduction thereof. Despite being highly cost-effective in terms of cost of treatment per child at scale, there are relatively few available studies reporting the overall costs involved in deworming at sufficient detail for efficiency analysis [7]. Of those available, even fewer have focused on the cost of large-scale surveying [7]. This is despite recent innovations in study design, which may allow for improvements in precision and, in turn, in cost-effectiveness over traditional survey methods [13]–[16]. The goal of this article is therefore to review the costs involved in conducting a school-based survey of STH and schistosomiasis and couple them with recent analysis of a novel surveying approach to compare the cost-effectiveness of different survey methodologies and to understand consequent cost-precision trade-offs.

### Survey approaches

The traditional approach to large-scale representative surveying of STH and often schistosomiasis has been to use two-stage random sampling or modifications thereof, often stratified by ecological zone [10], [12]. However, established literature suggests that the prevalence of helminth infection is highly predictable based on environmental variables, treatment frequency, and other geo-spatial correlates [13]–[16]. This suggests that traditional survey design estimates may be inefficient, because they only partially account for geographical variation [13]–[16]. This presents the opportunity for a more efficient survey design incorporating geospatial and other risk covariates, generally referred to as a model-based-geostatistical (MBG) approach [13]–[16].

Critically for the cost-effectiveness of helminth control programs, MBG differs from traditional survey design in the selection of sites for surveying and the information which is derived from those sites post-survey. Traditional design suggests randomization of sites for surveying across representative areas, such as ecological zones. MBG uses predictive models to identify sites which provide the most predictive power for post-survey modelling of prevalence. In this way, purposive sampling can be used to target the most informative sites and maximize survey precision under given resource constraints. In addition, traditional methods for post-survey estimation of prevalence do not consider prior prevalence, most risk factors, or geospatial variation. MBG, however, uses this information post-survey to create predictive models of prevalence which are as accurate as possible, under stated assumptions. Such targeting also maximizes the potential integration of STH and schistosomiasis, given their similarities in risk-factors [13]–[16].

### Kenyan National School Based Deworming Program

Kenya is endemic with both STH and schistosomiasis, and in 2012 Kenya launched the National School Based Deworming Program (NSBDP), providing PC to SAC and PSAC for both diseases in selected counties across the country [17]. As part of the NSBDP, a comprehensive monitoring and evaluation strategy was implemented by the Kenya Medical Research Institute (KEMRI), which included large-scale representative surveys using two-stage cluster randomization at baseline, year three, year five, and year six of the program.

## Methods

### Method overview

To identify the relative cost-precision of traditional surveying against MBG, retrospective micro-costing of the NSBDPs’ year five survey was performed, which covered both STH and schistosomiasis in 2017. Costing estimates were then paired to various precision estimates derived from a previously performed simulation exercise based on surveys conducted in Kenya between 2012 and 2017 [15]. This exercise involved deriving theoretical prevalence estimates in 2017 for both traditional surveying, which was used during the 2017 survey, and MBG, and comparing them with empirical results obtained from the 2017 survey. By comparing simulated estimates with empirical results a relative level of precision for each method was quantified. Results are presented as: costing and cost analysis of the 2017 survey; cost-precision of the surveying approaches under different budget constraints; and Incremental Cost Effectiveness Ratios (ICERs) of MBG compared to selected budget thresholds.

To analyze costs and compare surveying approaches under different budget scenarios, cost and precision metrics were simulated under combinations of the number of schools sampled, and the number of children surveyed per school. This process involved first compiling cost components from the original 2017 survey, then extrapolating these to show how costs differ when altering both variables. Each school/participant combination aligned with a precision estimate using traditional surveying and using MBG. Cost and precision were simulated for 36 total scenarios between traditional surveying and MBG. Aside from site selection, field procedures for conducting traditional surveying and MBG are identical. Therefore, the total cost by scenario was assumed to be identical between traditional and MBG survey designs that use the same number of schools and samples per school. To estimate ICERs four traditional surveying scenarios were selected to act as cost-effectiveness thresholds. These scenarios represent iterations of WHO recommended traditional surveying [10]. Each of the four scenarios was compared to all 36 MBG scenarios and placed across a cost-effectiveness plane. A conceptual framework of the study design is provided in figure 1 below. Sample numbers and traditional surveying reference cases are provided in table 1 below.

**Figure 1.**
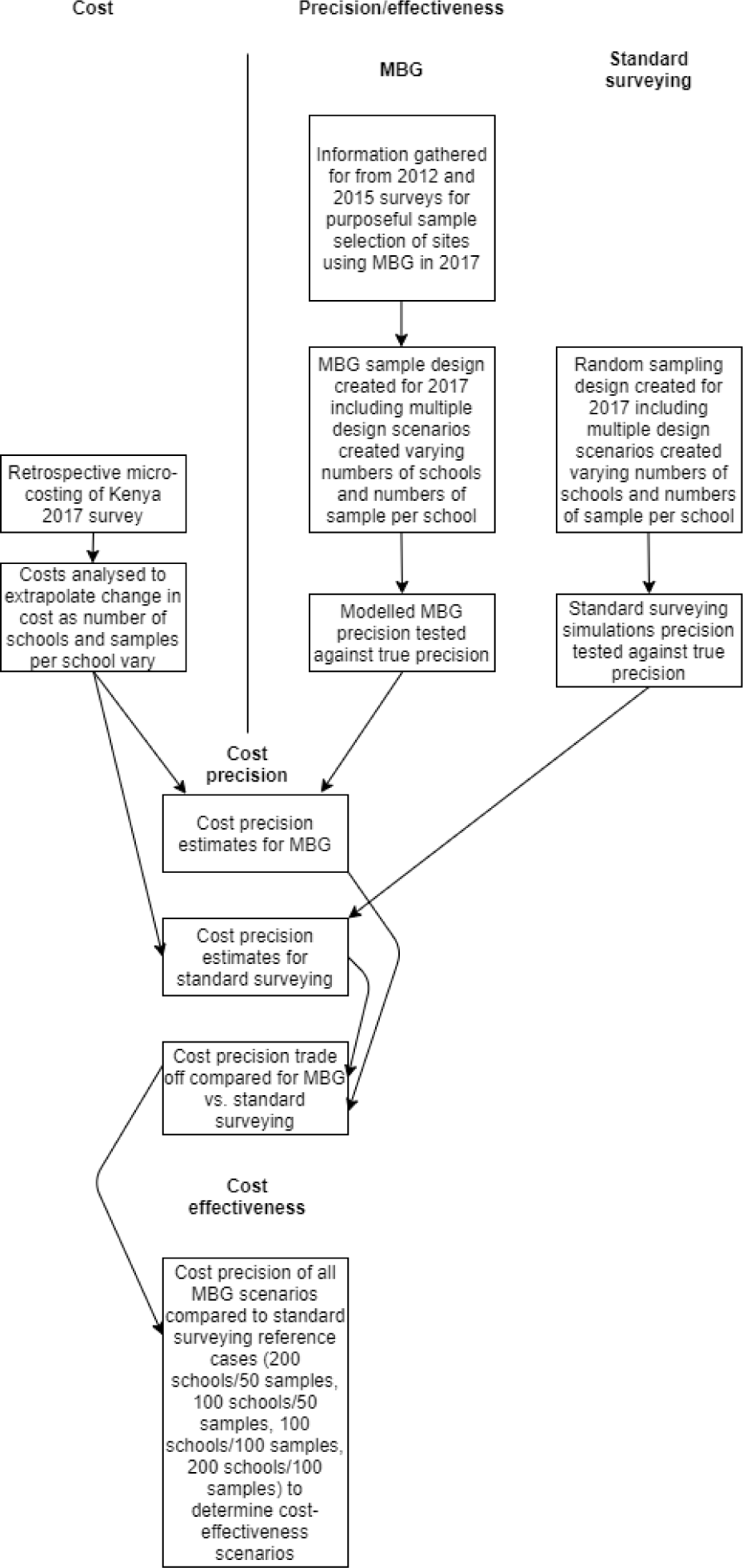
Conceptual framework of study design.

**Table 1.**
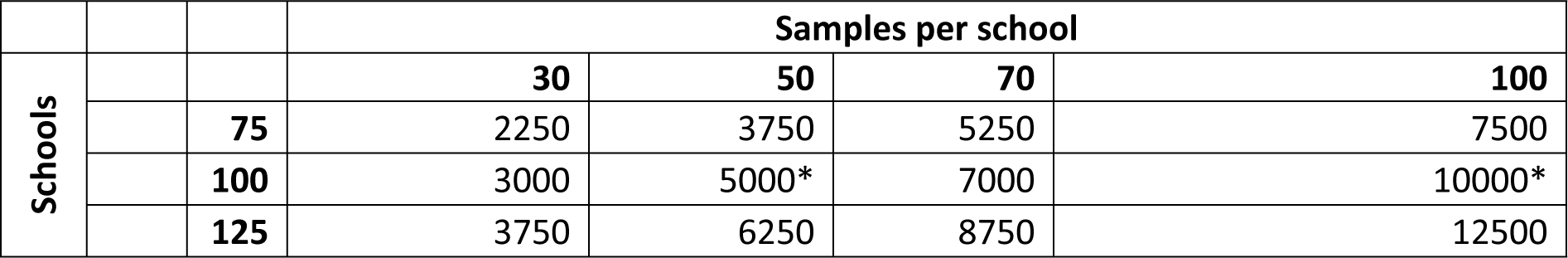
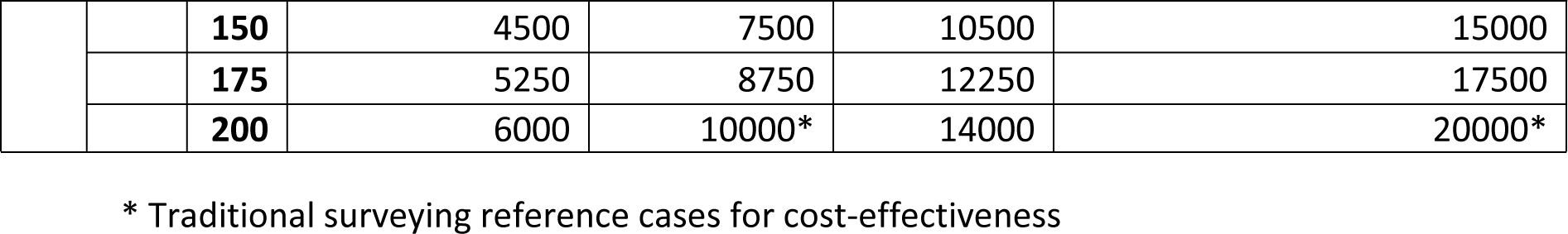
Sample design scenarios, number of schools, samples per schools and overall samples.

### Data

We utilize data gathered from the NSBDP in Kenya, which conducted three successive large-scale surveys of STH and schistosomiasis prevalence in 2012, 2015, and 2017 (with a fourth performed in 2018, not used in the present analysis). Details of the surveys and their methodology have been published elsewhere [15]. Costs are derived from expenditure generated during the 2017 survey. Simulations for relative precision of survey designs use data from 2012, 2015 and 2017 performed elsewhere [15]. Ethical approval for all surveys was obtained from the KEMRI’s scientific and ethical review committee for the original surveys (SSC Number 206), all surveys involved informed consent. All further analysis was conducted on de-identified secondary data and exempt from ethical review.

### Costing methods

Following methods outlined in Drummond et al [20], a retrospective compilation and analysis of costs and cost components was performed. Cost records were reviewed, and expert opinion of survey implementers was obtained regarding the frequency and specifics of survey costs. An ingredients-based approach was used to compile costs, reviewing individual cost components and apportioning them to cost categories. Financial costs only were estimated, from the perspectives of the survey donor and the Ministry of Health. As the primary purpose of the survey was to compare cost-effectiveness of survey designs in isolation, we excluded all economic costs and in-kind resources, such as opportunity cost of primary school teachers’ time, and principal investigator research time, including protocol development and analysis. To estimate the total value of goods or services, unit price was multiplied by the total number of each item. Total costs were extrapolated by survey scenario, school, and individual sampled to be able to capture the variation in cost-precision when modifying the school/sample size. Average costs per individual sampled, school sampled, and scenario were also calculated. An exchange rate of 92 Kenyan Shilling per U.S. dollar (USD) for the costs of survey activities between 2017 and 2022 was used [21].

### Traditional surveying

Traditional surveying approaches refer to current common practice in designing nationally representative surveys for STH and schistosomiasis to advise decisions on PC frequency. This involves two-stage random cluster sampling, with randomization performed at both the selection of individual schools (sites) and random selection of children within schools, however stratified across ecological zone [10]. The method simulated is identical to the first survey conducted in Kenya [19], with a modified version, incorporating sentinel surveillance, currently recommended by the WHO [10].

### Model-based geostatistical surveying

MBG methods use prior geospatial information to optimize site selection at the design stage, and exploit any spatial correlation in the resulting data to maximize precision under given resource constraints. For a complete account, including technical details of the MBG method, see Diggle and Giorgi (2019). Briefly, a prevalence survey conducted in a school at a geographical location *x* generates data in the form of a pair of values: *n*, the number of individuals tested; and y, the number positive. The sampling distribution of *y* is binomial with number of trials *n* and probability of positive outcome *P(x),* the school-wide prevalence at *x*. Variation in *P(x)* over the region of interest can be explained by a combination of three phenomena: context-specific covariate effects *d(x)* (for example, land surface temperature as a proxy for suitable breeding conditions for the responsible vector); unexplained residual spatial variation, *S(x),* which is represented in this case as a latent stochastic process; unexplained residual non-spatial variation, *Z*, which is represented as a set of independent and Normally distributed random effects.

The traditional way to account for these three 3 components is with a logistic geospatial model:

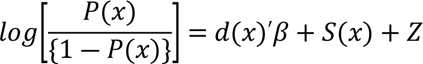

Crucially, whereas classical statistical methods ignore the stochastic term *S(x),* geospatial methods include it, estimate its properties from the data, and exploit its spatial correlation structure to give more precise predictions of *P(x).* Informally, the geospatial modelling pools information from neighboring locations, to an extent justified by the data, and so increases precision without sacrificing spatial resolution or inferential probity.

Spatially regulated sampling imposes a minimum distance between any two sampled locations. In the presence of spatial correlation, this usually leads to better predictive performance than spatially random sampling because it avoids sampling near-neighboring schools that essentially duplicate each other’s information. Given that MBG is able to make qualified predictions about future endemicity, sample site selection can be optimized to gain maximum information about the distribution of a given disease across a geography. In this way, site selection using MBG employs a spatially-regulated selection method.

### Effectiveness/precision metric

A precision metric was derived to determine relative effectiveness of comparative survey designs. Details of the simulation exercise used to derive the metric are provided elsewhere [15]. Briefly, to determine the relative precision of surveying approaches, predictive prevalence of both STH and schistosomiasis was modelled across all NSBDP counties in 2017, using 2017 survey data and other covariates. Simulations of study designs were then performed utilizing random sampling in the case of traditional surveying and spatially-regulated sampling based on 2012 and 2015 data in the case of MBG. Using the modelled simulations counties were classified into WHO recommended prevalence ranges [10]. Ranges were compared to the 2017 modelled prevalence surface data, and the correct proportion of counties correctly identified were obtained, creating a survey precision metric. The exercise was performed for STH and for schistosomiasis. Individual precision estimates for STH and schistosomiasis were averaged to obtain a single metric in view of the integrated surveying approach. Simulations for both designs were evaluated across varying scenarios of total schools and students sampled per school.

### Cost-precision and cost-effectiveness

To determine the relative changes in survey efficiency we couple cost and precision data for traditional surveying and MBG, across all sample size scenarios. This is presented as both cost per gain in precision, and difference in cost-precision between survey designs. To determine cost-effectiveness, four traditional surveying cost-precision estimates are used as reference cases and are compared to MBG scenarios to generate ICERs. Reference cases are selected to represent budget constraints that countries may be facing or survey sizes that have already been implemented.

## Results

### Costs

The overall cost of the year five NSBDP survey, when comprised of 200 schools with 100 students per school, is estimated to be $529,314 USD. This comprises an initial cost to reach the first school/sample including training, salary and transport costs for sufficient staff and equipment for sampling one participant of $26,000, with a marginal cost of $25 per additional sample or $2,516 per additional school. The largest proportion of costs are found to be lab equipment compromising 44.19% of total, followed by salary, and travel. The breakdown of costs by activity/item can be seen in table 2 below. A detailed description of components of cost activity/items can be found in annex x.

**Table 2.** Total cost of survey of 200 schools, 100 individuals per school.

| Category | Survey costs (200 schools, 100 samples) |  |
| --- | --- | --- |
|  | Cost (USD) | Proportion of overall cost |
| Salary | \$201,848 | 38.13% |
| Travel | \$9,275 | 1.75% |
| Venue hire | \$11,298 | 2.13% |
| Lab equipment | \$255,075 | 48.19% |
| Survey equipment | \$1,196 | 0.23% |
| Incidentals | \$50,623 | 9.56% |
| **Total** | **\$529,314** | **100.00%** |

Cost by survey scenario (**Figure 2**) are shown to increase with total sample size. The average cost per sample (**Figure 3**) was highest when minimizing both samples per schools and schools visited ($82.43 for 75 schools 30 samples per school, total 2250 samples), and lowest when maximizing both ($26.5 for 200 schools, 100 samples per school, total 20,000 samples) suggesting that the number of schools is a stronger driver of cost than the number of samples per school. A marginally decreasing cost minimization is found as samples per school are maximized. As an example, cost per sample for 75 schools/30 samples per school is $82.42, but for 200 schools/30 samples is $73.24, a difference of $9.18 per sample. When comparing 75 and 200 schools but with 100 samples per school, the cost difference is reduced to $2.76.

**Figure 2.**
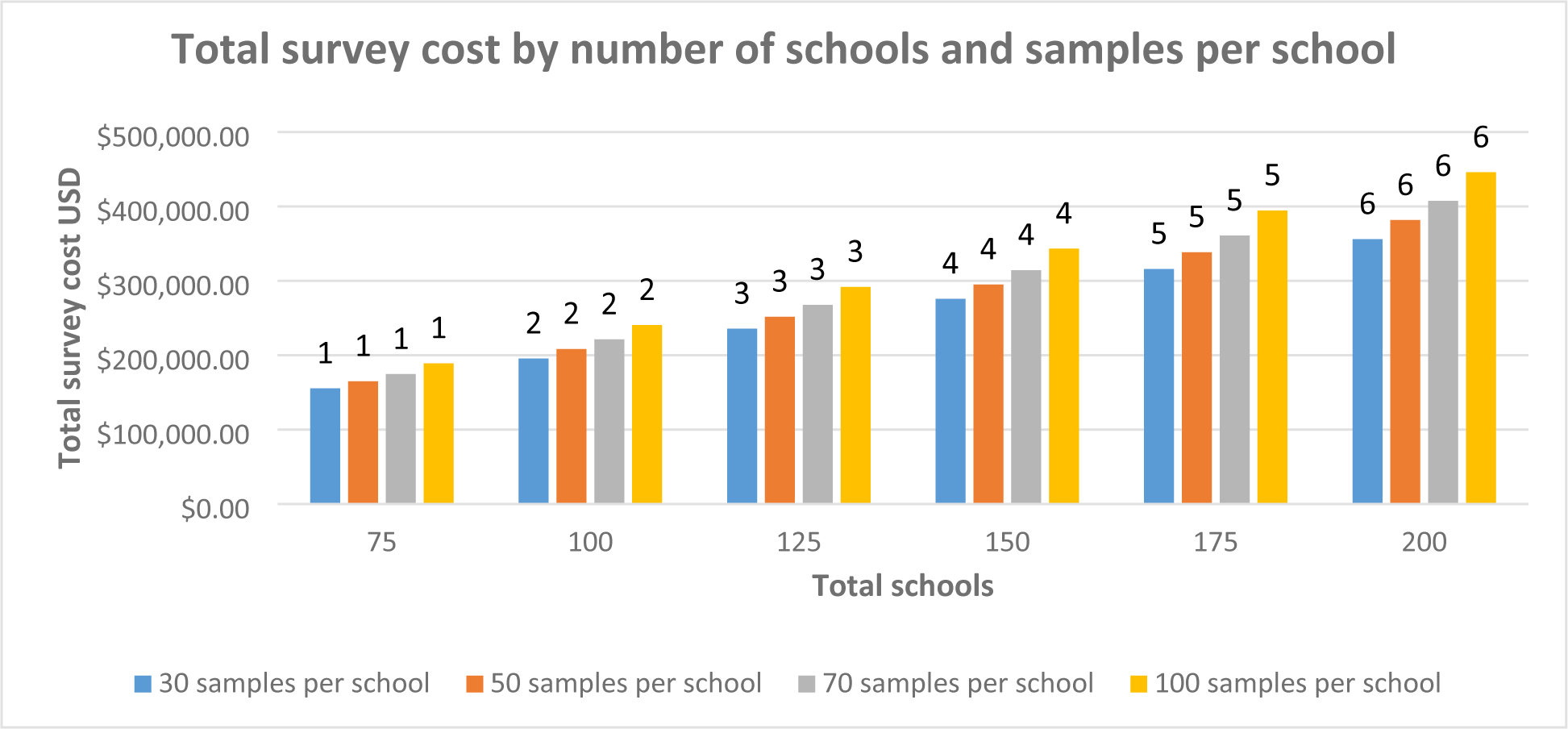
Total survey scenario cost by number of schools and samples per school.

**Figure 3.**
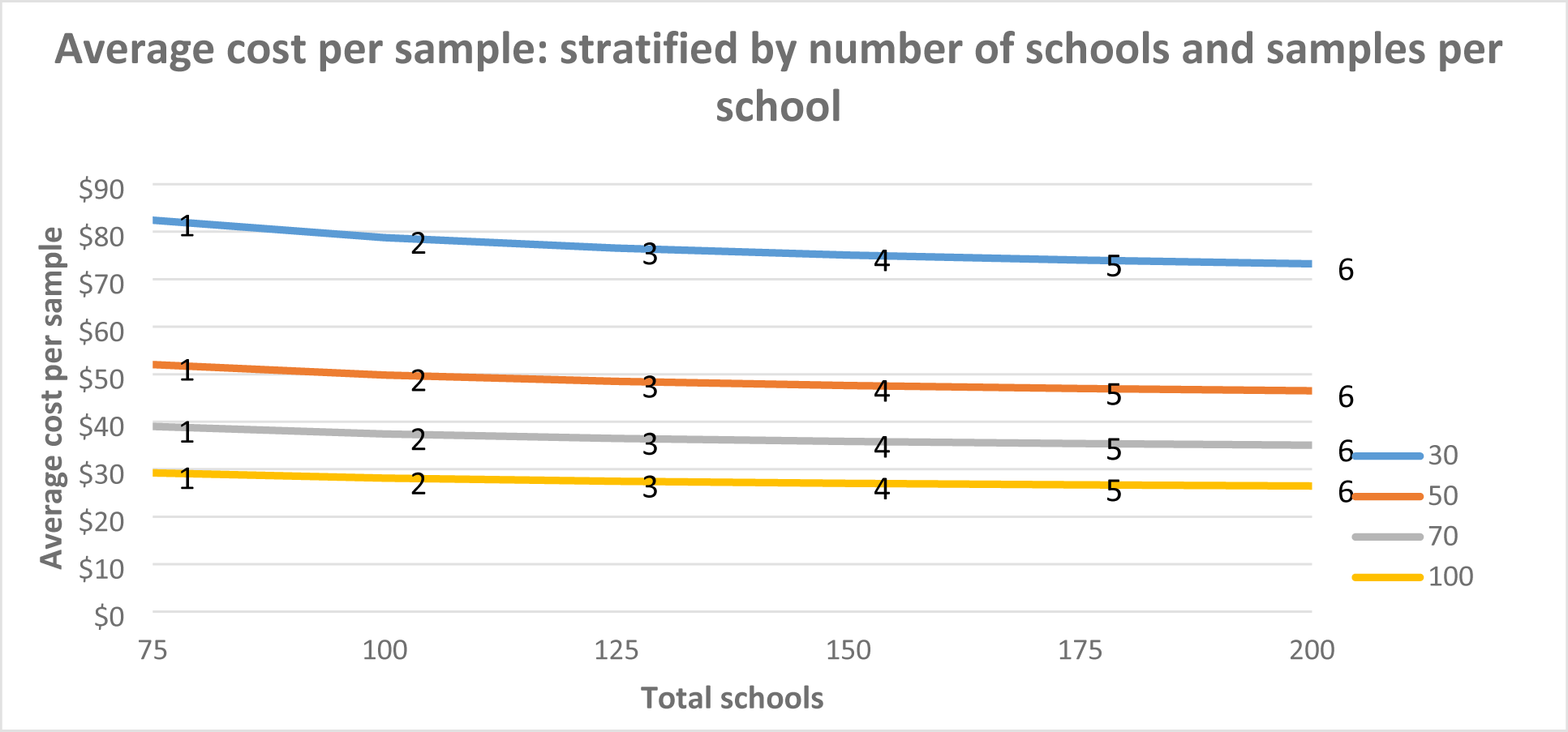
Average cost per sample by number of schools and samples per school.

Surveys covered sampling for both STH and schistosomiasis. There was no additional cost in training/set up, or in reaching schools between STH and schistosomiasis. Identification of STH and *S. mansoni* generally involve double-slide Kato Katz, with identification able to proceed using the same slides given appropriate training. *S. haematobium,* however, uses a urine filtration method which represents the primary cost difference when surveying STH and schistosomiasis. We observe the cost for adding both schistosomiasis species to an STH survey, assuming that equipment such as slides, forceps etc. is already included in STH costing. When surveying 200 schools with 100 samples per school, extra equipment for urine filtration costs a total of $21,900 overall for all schools. This reflects 4.13% of overall survey costs and equates to $109.48 per school or $1.09 per sample. It should be noted however, that in most situations, not all schools will sample for *S. haematobium* given the focal nature of the disease.

### Cost-precision

Results from the simulation exercise used to derive precision estimates [15] show that the MBG survey approach is more precise in correctly identifying county prevalence for both STH and schistosomiasis under all survey resource scenarios. Given that costs between the two design and analyses are identical given the same sample size, MBG is systematically more cost-precise. This relationship is conceptualized as the relative cost savings that can be achieved given a specific precision requirement as shown in Table 3 and Figure 4 below. The highest precision simulated under traditional design and analysis of 94.7% requires sampling 200 schools with 100 samples per school at a total cost of $446,000 USD. Comparable precision may be gained using MBG by surveying 200 schools/30 children ($356,000), 125 schools/70 children ($267,719), or 100 schools/100 children ($240,000).

**Figure 4.**
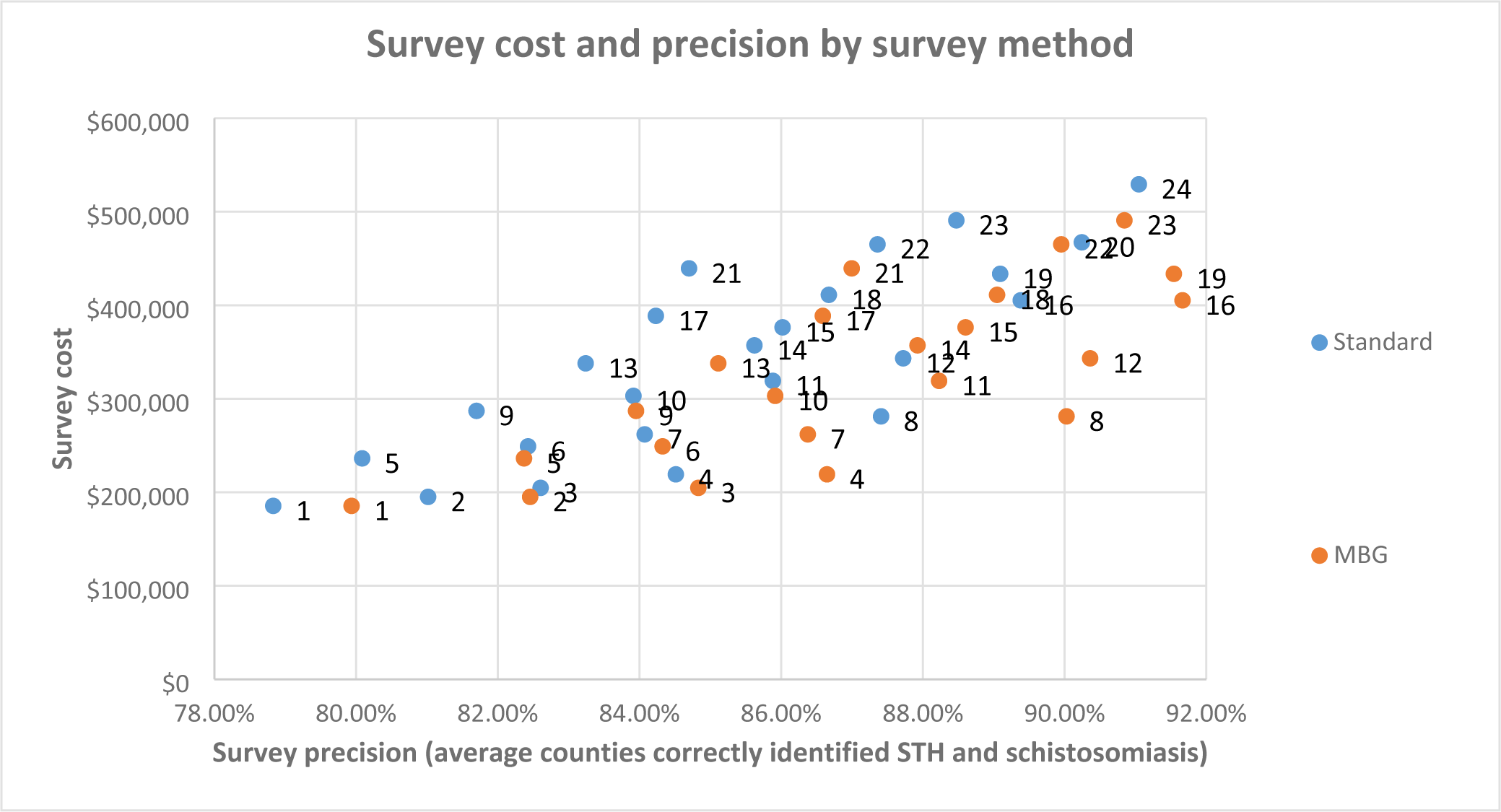
Cost and precision of survey scenario by survey design.

**Table 3.** Cost, precision, and cost per precision of survey designs.

| Sample scenario<br>(schools/<br>samples) | Costs |  |  | Traditional surveying |  | MBG |  | Cost difference<br>per precision<br>percentage |
| --- | --- | --- | --- | --- | --- | --- | --- | --- |
|  | Cost | Cost per<br>school | Cost per<br>sample | Average<br>precision<br>STH and<br>SCH | Cost per<br>1%<br>precision | Average<br>precision STH<br>and SCH | Cost per<br>precision<br>percentage |  |
| 75/30 | \$185,457 | \$2,473 | \$82 | 78.83% | \$2,353 | 79.94% | \$2,320 | \$33 |
| 75/50 | \$195,086 | \$2,601 | \$52 | 81.02% | \$2,408 | 82.46% | \$2,366 | \$42 |
| 75/70 | \$204,715 | \$2,730 | \$39 | 82.60% | \$2,478 | 84.83% | \$2,413 | \$65 |
| 75/100 | \$219,158 | \$2,922 | \$29 | 84.51% | \$2,593 | 86.65% | \$2,529 | \$64 |
| 100/30 | \$236,254 | \$2,363 | \$79 | 80.09% | \$2,950 | 82.37% | \$2,868 | \$82 |
| 100/50 | \$249,093 | \$2,491 | \$50 | 82.43% | \$3,022 | 84.33% | \$2,954 | \$68 |
| 100/70 | \$261,931 | \$2,619 | \$37 | 84.07% | \$3,116 | 86.38% | \$3,032 | \$83 |
| 100/100 | \$281,189 | \$2,812 | \$28 | 87.41% | \$3,217 | 90.03% | \$3,123 | \$94 |
| 125/30 | \$287,052 | \$2,296 | \$77 | 81.70% | \$3,514 | 83.95% | \$3,419 | \$94 |
| 125/50 | \$303,100 | \$2,425 | \$48 | 83.91% | \$3,612 | 85.92% | \$3,528 | \$84 |
| 125/70 | \$319,148 | \$2,553 | \$36 | 85.88% | \$3,716 | 88.23% | \$3,617 | \$99 |
| 125/100 | \$343,221 | \$2,746 | \$27 | 87.72% | \$3,913 | 90.36% | \$3,798 | \$114 |
| 150/30 | \$337,849 | \$2,252 | \$75 | 83.24% | \$4,059 | 85.11% | \$3,969 | \$89 |
| 150/50 | \$357,107 | \$2,381 | \$48 | 85.62% | \$4,171 | 87.92% | \$4,062 | \$109 |
| 150/70 | \$376,365 | \$2,509 | \$36 | 86.02% | \$4,375 | 88.60% | \$4,248 | \$128 |
| 150/100 | \$405,252 | \$2,702 | \$27 | 89.38% | \$4,534 | 91.66% | \$4,421 | \$113 |
| 175/30 | \$388,646 | \$2,221 | \$74 | 84.23% | \$4,614 | 86.59% | \$4,488 | \$126 |
| 175/50 | \$411,114 | \$2,349 | \$47 | 86.67% | \$4,743 | 89.05% | \$4,617 | \$126 |
| 175/70 | \$433,581 | \$2,478 | \$35 | 89.09% | \$4,867 | 91.54% | \$4,736 | \$130 |
| 175/100 | \$467,283 | \$2,670 | \$27 | 90.24% | \$5,178 | 92.43% | \$5,056 | \$122 |
| 200/30 | \$439,444 | \$2,197 | \$73 | 84.70% | \$5,188 | 87.00% | \$5,051 | \$137 |
| 200/50 | \$465,121 | \$2,326 | \$47 | 87.36% | \$5,324 | 89.95% | \$5,171 | \$154 |
| 200/70 | \$490,798 | \$2,454 | \$35 | 88.47% | \$5,547 | 90.84% | \$5,403 | \$145 |
| 200/100 | \$529,314 | \$2,647 | \$26 | 91.05% | \$5,814 | 93.03% | \$5,690 | \$124 |

### Incremental cost-effectiveness of MBG

ICERs of all MBG scenarios against four traditional design reference cases (schools/samples per school; 200/100; 200/50; 100/100; 100/50) can be seen across cost-effectiveness planes below in Figures 5 and 6, while absolute values are presented in Table 4 below.

**Figure 5.**
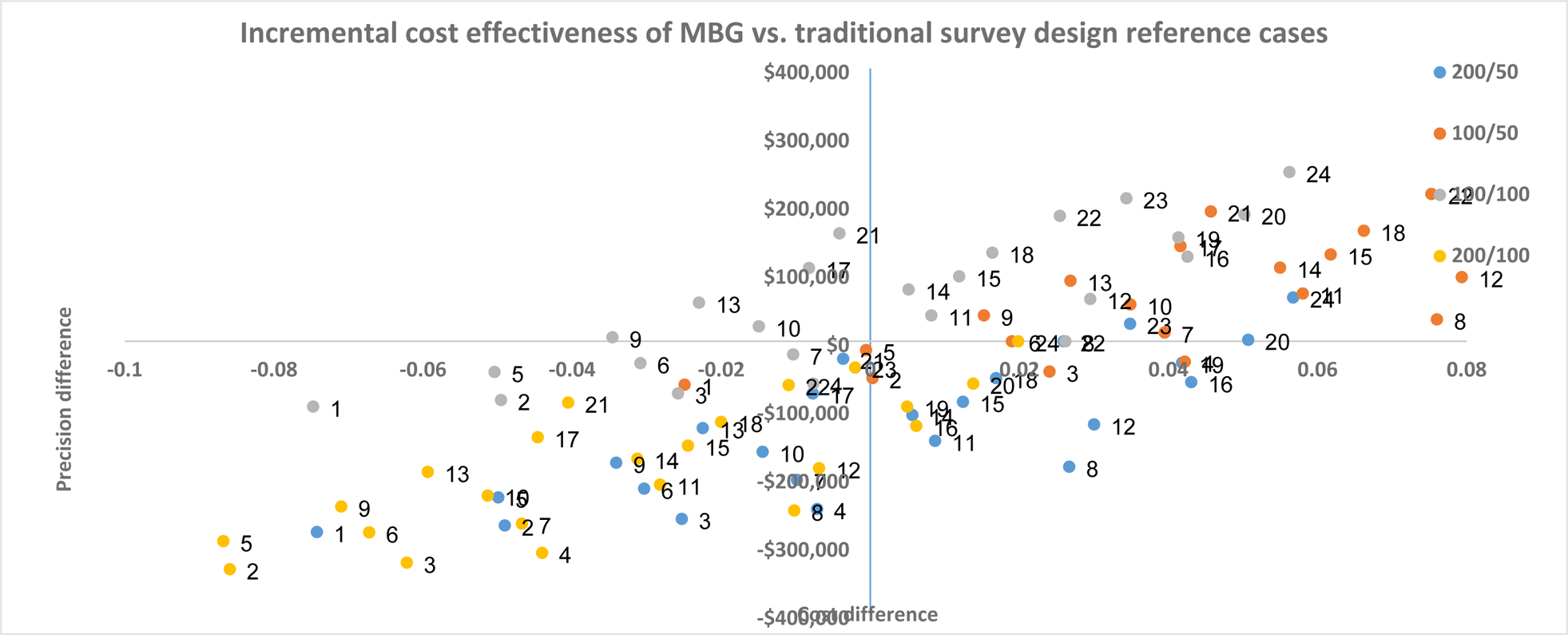
Incremental cost effectiveness of MBG vs. traditional survey design across all reference cases.

**Figure 6.**
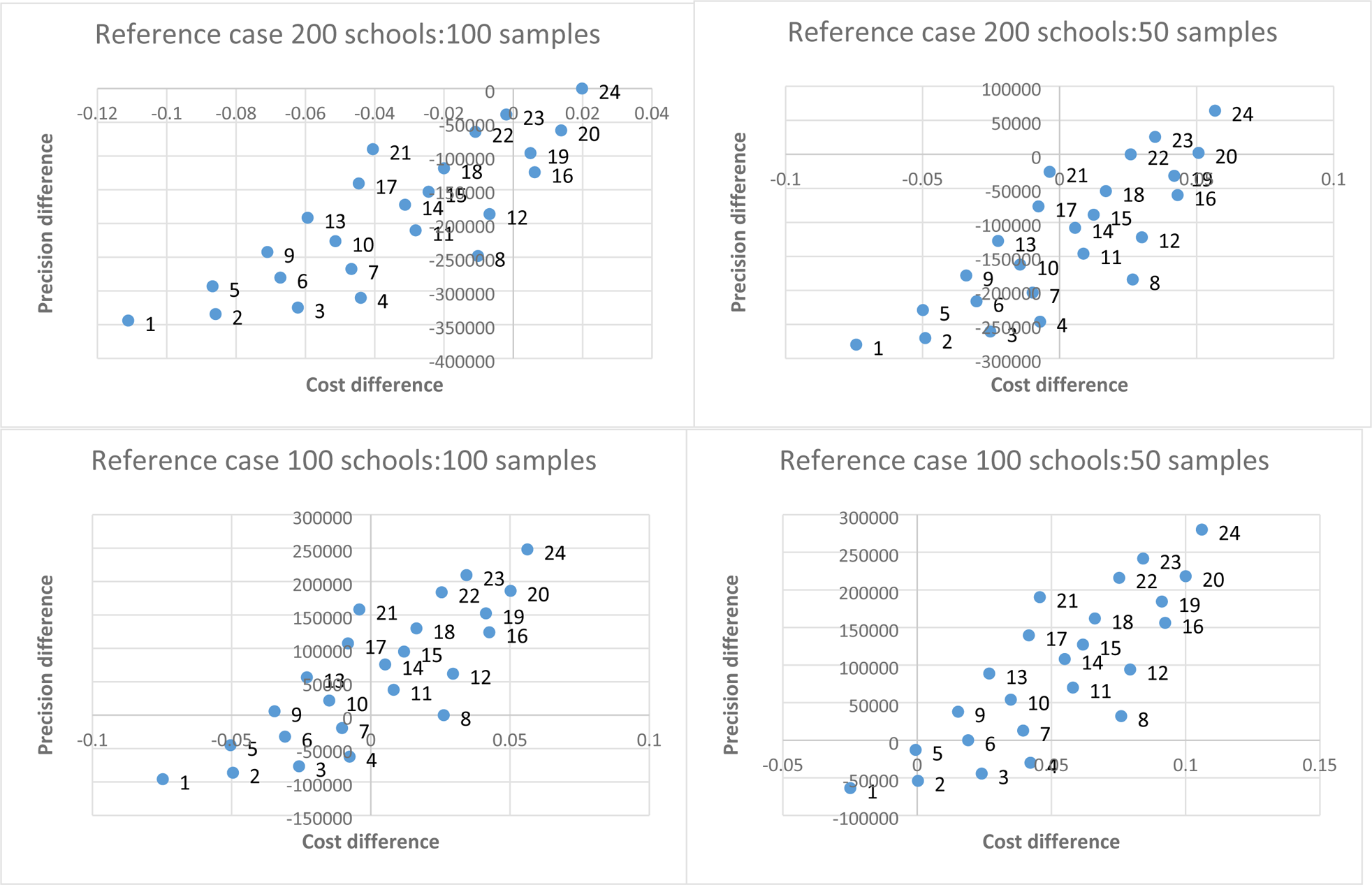
Incremental cost effectiveness of MBG vs. traditional survey design by reference case.

**Table 4.** Difference in cost and precision between traditional surveying reference cases and MBG scenarios.

|  | Reference case<br>200schools:100 samples |  | Reference case 200<br>schools:50 samples |  | Reference case 100<br>schools:100 samples |  | Reference case 100<br>schools:50 samples |  |
| --- | --- | --- | --- | --- | --- | --- | --- | --- |
|  | Cost<br>difference | Precision<br>difference | Cost<br>difference | Precision<br>difference | Cost<br>difference | Precision<br>difference | Cost<br>difference | Precision<br>difference |
| **75/30** | -\$343,857 | -0.1111 | -\$279,664 | -0.0742 | -\$95,732 | -0.0747 | -\$63,636 | -0.0249 |
| **75/50** | -\$334,228 | -0.0859 | -\$270,035 | -0.0490 | -\$86,104 | -0.0495 | -\$54,007 | 0.0003 |
| **75/70** | -\$324,599 | -0.0622 | -\$260,406 | -0.0253 | -\$76,475 | -0.0258 | -\$44,378 | 0.0240 |
| **75/100** | -\$310,156 | -0.0440 | -\$245,963 | -0.0071 | -\$62,031 | -0.0076 | -\$29,935 | 0.0422 |
| **100/30** | -\$293,060 | -0.0868 | -\$228,867 | -0.0499 | -\$44,935 | -0.0504 | -\$12,839 | -0.0006 |
| **100/50** | -\$280,221 | -0.0672 | -\$216,028 | -0.0303 | -\$32,097 | -0.0308 | \$0 | 0.0190 |
| **100/70** | -\$267,382 | -0.0467 | -\$203,189 | -0.0098 | -\$19,258 | -0.0103 | \$12,839 | 0.0395 |
| **100/100** | -\$248,125 | -0.0102 | -\$183,932 | 0.0267 | \$0 | 0.0262 | \$32,097 | 0.0760 |
| **125/30** | -\$242,262 | -0.0710 | -\$178,069 | -0.0341 | \$5,862 | -0.0346 | \$37,959 | 0.0152 |
| **125/50** | -\$226,214 | -0.0513 | -\$162,021 | -0.0144 | \$21,911 | -0.0149 | \$54,007 | 0.0349 |
| **125/70** | -\$210,166 | -0.0282 | -\$145,973 | 0.0087 | \$37,959 | 0.0082 | \$70,055 | 0.0580 |
| **125/100** | -\$186,093 | -0.0069 | -\$121,900 | 0.0300 | \$62,031 | 0.0295 | \$94,128 | 0.0793 |
| **150/30** | -\$191,465 | -0.0594 | -\$127,272 | -0.0225 | \$56,660 | -0.0230 | \$88,756 | 0.0269 |
| **150/50** | -\$172,207 | -0.0312 | -\$108,014 | 0.0056 | \$75,918 | 0.0051 | \$108,014 | 0.0550 |
| **150/70** | -\$152,949 | -0.0245 | -\$88,756 | 0.0124 | \$95,175 | 0.0119 | \$127,272 | 0.0618 |
| **150/100** | -\$124,062 | 0.0062 | -\$59,869 | 0.0431 | \$124,062 | 0.0425 | \$156,159 | 0.0924 |
| **175/30** | -\$140,668 | -0.0446 | -\$76,475 | -0.0077 | \$107,457 | -0.0082 | \$139,554 | 0.0416 |
| **175/50** | -\$118,200 | -0.0200 | -\$54,007 | 0.0169 | \$129,925 | 0.0164 | \$162,021 | 0.0662 |
| **175/70** | -\$95,732 | 0.0049 | -\$31,539 | 0.0418 | \$152,392 | 0.0413 | \$184,489 | 0.0912 |
| **175/100** | -\$62,031 | 0.0138 | \$2,162 | 0.0507 | \$186,093 | 0.0502 | \$218,190 | 0.1000 |
| **200/30** | -\$89,870 | -0.0405 | -\$25,677 | -0.0036 | \$158,254 | -0.0041 | \$190,351 | 0.0457 |
| **200/50** | -\$64,193 | -0.0109 | \$0 | 0.0259 | \$183,932 | 0.0254 | \$216,028 | 0.0753 |
| **200/70** | -\$38,516 | -0.0020 | \$25,677 | 0.0349 | \$209,609 | 0.0343 | \$241,705 | 0.0842 |
| **200/100** | \$0 | 0.0198 | \$64,193 | 0.0567 | \$248,125 | 0.0562 | \$280,221 | 0.1060 |

Cost per school/sample is shown to be identical between the two survey methods, while MBG is systematically more precise with the same resources. Consequently, under 87 of 92 school/sample reference case comparisons: MBG is cheaper with a precision trade off; more expensive with gains in precision; or both cheaper and more precise when compared to the traditional sampling reference cases. The thirteen comparisons in which MBG is both cheaper and more precise are found when reference cases are 200 schools/100 samples, 200 schools/50 samples, or 100 schools/50 samples. Five instances are noted where MBG is more expensive and less precise, all when the reference case is 100 schools/100 samples, compared to MBG scenarios which utilize 30 samples per school, and in one case 50. These cases represent extremes of simulations used to evaluate MBG and are unlikely to be utilized in practice.

## Discussion

When compared with traditional surveying, our analysis shows MBG to be more cost-effective under 95% of school/sample size reference case comparisons. The use of reference cases is designed to compare the relative benefit of MBG to traditional surveying approaches/budgets already implemented. A high frequency of nationally representative surveying of STH has employed sample sizes calculated using one school per 200,000-300,000 SAC with 50 students selected per school, as per current WHO guidelines [10]. In all comparisons using reference cases with 50 students selected per school, MBG is more cost-effective -- maximizing precision, minimizing cost, or in many cases both. Importantly, as the primary difference between the two methods is the use of random or spatially-regulated site selection and analysis, survey cost does not differ between the two methods when surveying identical numbers of schools and study participants. For identical cost, however, MBG is always more precise. Consequently, MBG will be either more effective for the same cost, or cost-saving when requiring the same precision regardless of the sample size selected when compared to survey approaches currently commonly implemented.

In settings where helminth prevalence has been reduced to very low levels, high frequency PC is decreasingly cost-effective due to the ratio of children needing treatment and those receiving it. At 2% prevalence, only two out of every 100 children will be infected, yet the entire cohort will be targeted to receive deworming medication. While it is critical that all children with helminths be attended to, subsequent deworming of an entire population becomes less cost-effective relative to other interventions and programs within national health budgets. WHO recommendations suggest reducing treatment frequencies in line with prevalence reductions, including suspension of PC, but with continued monitoring where STH or schistosomiasis prevalence is less than 2%. This is to ensure that resurgence is avoided when treatment suspension is warranted [22]. To maintain the cost-effectiveness of deworming, novel surveying strategies and avenues for maximizing cost-precision as a critical component of long-term deworming strategy should be explored. This is to ensure that deworming remains within health budgets and years of steady progress in elimination of both STH and schistosomiasis as a public health problem not be lost [22].

Our costing analysis shows more than 40% of survey expenses are driven by lab equipment. Consumables used for diagnosis include the equipment used for the Kato Katz method, which is recommended for determining population-level prevalence [10], and is currently the most cost-effective diagnostic method for surveying in most situations [23]. Cost-minimization of diagnostics has been explored elsewhere [23] and alternate diagnostic methods are available or in development. No method, however, has yet been shown as sufficiently cheaper per diagnosis to warrant adoption as traditional, particularly in the case of STH. Some methods, such as semi-quantitative PCR have focused on improving precision, particularly at low levels of prevalence, and less on ensuring cost-efficiency for individual diagnoses [24]. A full economic evaluation of PC including long-term scenarios such as parasite elimination may estimate these techniques to be more cost-effective, particularly when considering the need for confirmation of elimination and sample batching. However, at present, the path towards elimination of both STH and schistosomiasis remains unclear and, in the immediate to medium term, Kato Katz offers the greatest cost-precision for elimination as a public health problem. Methods for reducing surveyor time are also becoming available, such as machine learning based recognition of slides [25]. However, efficiency gains from these developments are likely to be limited, as sample collection remains necessary for diagnosis. At present, efficiency gains, highlighted in our study show site selection and MBG to be important avenues for exploring continued sustainability of deworming, particularly as it enters a new era focused on maintaining cost-effectiveness.

Our analysis highlights three avenues of maximizing cost-precision gains using MBG when surveying STH and schistosomiasis. Firstly, return on initial investments can be maximized by exploiting economies of scale due to fixed and step-fixed costs. We estimate that approximately $26,000 USD is spent to reach the first school, comprising 14% of total costs when considering a survey of 75/30 schools/samples and only 5% when considering 200/100 schools/samples. These upfront costs remain fixed as they primarily involve field staff training. Consequently, the more samples taken, the more efficient the single upfront investment becomes. Upon reaching a school, a step-fixed cost occurs, as number of samples can be increased to maximize efficiency of the cost of travel and surveyor time. As implied by the name, this cost increases at stepped intervals, as total sample number has an upper limit per school under fixed resources and additional samples beyond this point require hiring of more personnel and paying for their travel. Secondly, by using MBG, cost-precision between schools and samples per school can be optimized. Both fixed and step-fixed costs experience a decreasing marginal cost saving per sample, along with a marginally decreasing gain of precision. Any design calculation is based on a working model for the data to be collected. MBG uses historic data and context-specific information, for example on recent treatment history, to construct a working model of current prevalence and uses this model to estimate which sites are best able to deliver precision. Given an available estimate of precision, relative costs of school and sample number can be mathematically optimized pre-survey to yield the most cost-precise estimates available given a required budget or accuracy. Site and sample optimization has been explored prior for traditional surveying [26] however only as a generic modelling exercise for all geographies, with precision gains subject to chance due to the necessary randomization of sites. This is demonstrated through the improved precision and cost-effectiveness seen within our analysis. Thirdly, economies of scope can be used to maximize cost-efficiency through integrated surveying. Both STH and schistosomiasis share similar requirements for sampling, along with risk factors for higher prevalence. In many cases this may be limited by the specific schistosomiasis species being surveyed, as *S. mansoni and S. haematobium* require different diagnostic techniques. Additionally, in some cases mapping an entire geography for schistosomiasis may not be cost-effective given the highly focal nature of the parasite [23].

Several limitations of our study are noted. Firstly, all results are modelled post-hoc using available data. Precision estimates are gained using simulations based on real world data; however, it is unlikely that a valid counterfactual can ever be derived for comparing cost-effectiveness of representative cost-effective surveying. Secondly, results obtained from this exercise may not be directly transferrable to other geographies. The present study relies heavily on data obtained from deworming in Kenya which may differ in cost and availability of survey resources and geographic variation of STH and schistosomiasis risk. Thirdly, cost components measured during this study focused primarily on those involved in the direct surveying component of prevalence measurement. Knowledge of geostatistical methods is unlikely to be universal across data analysts working with NTDs, and as such would likely require some level of training, which adds additional costs when comparing survey designs. Such costs however, are expected to be minimal and occur only once.

## Conclusion

The transmission of STH and schistosomiasis are influenced by environmental and socioeconomic factors, along with rounds of PC, making prevalence and intensity highly predictable comparative to other infectious diseases. Predictability of prevalence allows improved precision to be incorporated into survey designs. Our analysis demonstrates that the predictability of both diseases can be exploited to gain cost-efficiency when surveying. A survey design incorporating such predictability through the use of geostatistical methods will often be preferential over traditional survey methods for STH and schistosomiasis through improved precision for identical costs or reduced costs for similar levels of precision. Optimizing survey design will become increasingly important as prevalence decreases to ensure that deworming remains a priority within national health budgets and years of steady progress are not lost.

## Data Availability

This work combines two sections of data. The original simulation data can be found in pre-existing publications. Detailed costing data has been provided as supplementary material as part of this manuscript.

---

**Supplementary material 1:** Itemized cost for individual survey components, totaled for a survey comprising of 200 schools and 100 students per school.

| Stage | Activity/item class 1 | Item class 2 | Type or description | Total USD schools (200) samples (100) | Notes |
| --- | --- | --- | --- | --- | --- |
| Base-training | Travel | Training travel reimbursement | Travel + incidentals | 5980 | Fixed cost |
| Base-training | Venue hire | Conference hall hire | Single payment | 1104 | Fixed cost |
| Base-training | Lab equipment | Equipment hire | One off (per day) | 9200 | Fixed cost |
| Base-training | Venue hire | Lab space | One off (per day) | 920 | Fixed cost |
| Base-training | Salary | Lab trainers | Allowance | 1840 | Fixed cost |
| Base-training | Salary | Trainers' salary | Daily salary | 1840 | Fixed cost |
| Base-training | Incidentals | Data and process trainer allowance | Allowance | 920 | Fixed cost |
| Base-training | Salary | Data and process trainer salary | Daily salary (in kind) | 1840 | Fixed cost |
| Base-training | Venue hire | Hire of training facility | Rental | 73.6 | Fixed cost |
| Base-training | Incidentals | Refreshments + lunch | Food/beverage | 23 | Fixed cost |
| Base-training | Lab equipment | Consumables (training) | Disposable equipment | 460 | Fixed cost |
| Sampling | Travel | Fuel costs - near | Average | 2906.28 | Multiplied per school or per sample |
| Sampling | Travel | Fuel costs - far | Average | 388.4608 | Multiplied per school or per sample |
| Sampling | Salary | Field staff wage | Field supervisor | 50232 | Multiplied per school or per sample |
| Sampling | Salary | Field staff wage | Laboratory technologist | 33488 | Multiplied per school or per sample |
| Sampling | Salary | Field staff wage | Laboratory assistant* often local | 19320 | Multiplied per school or per sample |
| Sampling | Salary | Field staff wage | Data manager | 41952 | Multiplied per school or per sample |
| Sampling | Salary | Field staff wage | Data clerk | 19320 | Multiplied per school or per sample |
| Sampling | Salary | Field staff wage | Driver | 19320 | Multiplied per school or per sample |
| Sampling | Salary | Field staff wage | Cleaner | 11592 | Multiplied per school or per sample |
| Sampling | Salary | Local security | For security risk areas | 1104 | Multiplied per school or per sample |
| Sampling | Venue hire | Laboratory space | Space hire | 9200 | Multiplied per school or per sample |
| Sampling | Incidentals | Community mobilization materials | Per school, multiple days | 49680 | Multiplied per school or per sample |
| Sampling | Lab equipment | Labels | Packet (400 labels) 1 per school | 506 | Multiplied per school or per sample |
| Sampling | Lab equipment | Gloves | Packet (100) 2 schools | 1380 | Multiplied per school or per sample |
| Sampling | Lab equipment | Slide box | 2.5 boxes per school (100 slides) per school | 2208 | Multiplied per school or per sample |
| Sampling | Lab equipment | Cooler packs | 8 cooler boxes (used for all schools) 2 Per school (1 big and 1 small) | 588.8 | Multiplied per school or per sample |
| Sampling | Lab equipment | Cooler packs |  | 368 | Multiplied per school or per sample |
| Sampling | Lab equipment | Microscope | 2 needed per team (re-usable) (school) | 7360 | Multiplied per school or per sample |
| Sampling | Lab equipment | Forceps | 2 per team (re-usable) | 18.4 | Multiplied per school or per sample |
| Sampling | Lab equipment | Marker pens - pkts | 2 pens per school | 368 | Multiplied per school or per sample |
| Sampling | Lab equipment | Glycerol(liters) | 5 liters enough for whole exercise | 46 | Multiplied per school or per sample |
| Sampling | Lab equipment | Malachite green | 1 packet (100 grams) enough for whole exercise | 23 | Multiplied per school or per sample |
| Sampling | Lab equipment | Templates | 100 pieces per school | 92000 | Multiplied per school or per sample |
| Sampling | Lab equipment | Stool sieves | 50 pieces per school | 92000 | Multiplied per school or per sample |
| Sampling | Lab equipment | Buckets | 2 per school | 368 | Multiplied per school or per sample |
| Sampling | Lab equipment | Basin | 2 per school | 368 | Multiplied per school or per sample |
| Sampling | Lab equipment | Tally counters | 2 per school | 3680 | Multiplied per school or per sample |
| Sampling | Lab equipment | Sharps Containers | 1 per school | 920 | Multiplied per school or per sample |
| Sampling | Lab equipment | Slides: stool(100.box) | 2 packets per school (100 per packet) | 1840 | Multiplied per school or per sample |
| Sampling | Lab equipment | Stool poly pots(200/pkt) | 1 packet per school | 9200 | Multiplied per school or per sample |
| Sampling | Lab equipment | Wooden spatula 100/box | 2 packets per school | 736 | Multiplied per school or per sample |
| Sampling | Lab equipment | Slides Urine - 100/box | 2 packets per school | 736 | Multiplied per school or per sample |
| Sampling | Lab equipment | Urine chambers | 1 packet per school (100 chambers) | 4600 | Multiplied per school or per sample |
| Sampling | Lab equipment | Filters boxes of 100 | 1 packet per school (100 filters) | 3680 | Multiplied per school or per sample |
| Sampling | Lab equipment | Syringes - boxes | 100 pieces per school | 3680 | Multiplied per school or per sample |
| Sampling | Lab equipment | Urine containers | 1 packet per school (about 200 pieces) | 9200 | Multiplied per school or per sample |
| Sampling | Lab equipment | Toilet paper - bales | 5 pieces per school | 460 | Multiplied per school or per sample |
| Sampling | Lab equipment | Disinfectant:5litres | 10 liters for whole exercise | 3680 | Multiplied per school or per sample |
| Sampling | Lab equipment | Hand soap (Dettol) | 1 unit per school | 92 | Multiplied per school or per sample |
| Sampling | Lab equipment | Detergent(1kg) | 0.5 kg per school | 276 | Multiplied per school or per sample |
| Sampling | Lab equipment | Towels | 2 per school | 1472 | Multiplied per school or per sample |
| Sampling | Lab equipment | Toothbrush | 2 pieces per school | 184 | Multiplied per school or per sample |
| Sampling | Lab equipment | Biohazard bags | 1 piece per school | 184 | Multiplied per school or per sample |
| Sampling | Lab equipment | Bin bags | 1 piece per school | 147.2 | Multiplied per school or per sample |
| Sampling | Lab equipment | Bins | 1 piece per school | 920 | Multiplied per school or per sample |
| Sampling | Lab equipment | Newspaper | 0.5 kg per school | 46 | Multiplied per school or per sample |
| Sampling | Lab equipment | Paper towels - rolls | 1 roll per school | 276 | Multiplied per school or per sample |
| Sampling | Lab equipment | Biros(box) | 2 pens per school | 73.6 | Multiplied per school or per sample |
| Sampling | Lab equipment | Pencils(box) | 2 pencil per school | 73.6 | Multiplied per school or per sample |
| Sampling | Lab equipment | Lab coats (personal protective equipment) | 10 pieces for survey | 184 | Multiplied per school or per sample |
| Sampling | Lab equipment | Sanitizers | 0.5 liter per school | 552 | Multiplied per school or per sample |
| Sampling | Lab equipment | Masks | 1 box per school | 920 | Multiplied per school or per sample |
| Sampling | Survey equipment | Mobile phone | 1 per team | 276 | Multiplied per school or per sample |
| Sampling | Survey equipment | Mobile phone data (bundles) | 1000 mb per school | 920 | Multiplied per school or per sample |

